# Safety, efficacy and feasibility of preventive treatment for drug-resistant tuberculosis with moxifloxacin or bedaquiline

**DOI:** 10.1101/2025.03.09.25323266

**Authors:** Alexandra V. Solovyeva, Grigory V. Volchenkov, Oksana I. Ponomarenko, Tatiana A. Kuznezova, Tatiana R. Somova, Evgenia V. Belova, Sven Gudmund Hinderaker, Einar Heldal, Salmaan Keshavjee

**Author notes:** **Corresponding author:** Salmaan Keshavjee,; 641 Huntington Avenue, Boston, Massachusetts 02115.

## Abstract

**Introduction:** Rates of drug-resistant tuberculosis (TB) are increasing worldwide. TB preventive treatment (TPT) for contacts of active TB patients is essential to halt infection progression and transmission. While newer TPT regimens for drug-sensitive strains are expanding, optimal treatment for contacts exposed to drug-resistant TB (DR-TB) remains unclear. In 2019-2020, Vladimir City, Russia, introduced moxifloxacin and bedaquiline-based TPT regimens to prevent disease development in contacts exposed to DR-TB.

**Methods:** We conducted a retrospective cohort study using medical records data that included adult TB contacts, people experiencing homelessness, and persons with HIV who received TPT in Vladimir City, Russia, between 2019 and 2020. Those without TB disease but with indications for TPT were offered one of six regimens, based on drug-susceptible testing results of index patient: Rifapentine/Isoniazid (3HP), Isoniazid (6H), Rifabutin/Isoniazid (3HRb), Rifampicin (4R), Moxifloxacin (4Mfx), or Bedaquiline (3Bdq). Adverse drug reactions (ADRs) were monitored with monthly lab tests and ECGs.

**Results:** Over 24 months, 403 people started TPT. No life-threatening ADRs or deaths occurred. The lowest ADR rate and significantly higher completion rate was observed in 3Bdq (n=20, 95.2%) compared to 3HP (n=192, 75.9%, *Mid-P exact* = .03). The rate of TB disease per 1,000 person-years of observation was four times higher in individuals eligible for TPT who did not start it compared to those who initiated TPT.

**Conclusion:** Treatment for the prevention of DR-TB, including forms resistant to rifampicin and fluoroquinolones, is feasible, effective and safe. This study introduces a novel paradigm for TB prevention in high-burden DR-TB settings, offering a promising strategy to protect contacts and reduce transmission.

**Key points:** Moxifloxacin and bedaquiline are safe, effective, and feasible agents for preventive therapy among contacts of individuals with drug-resistant tuberculosis (TB) and can be used as part of the comprehensive search-treat-prevent approach for TB elimination.

## BACKGROUND

Individuals exposed to people with active tuberculosis may be infected with *M. tuberculosis* and develop TB infection (TBI; previously known as latent TB infection). Infection can progress to clinically active tuberculosis, but this can be prevented by tuberculosis preventive treatment (TPT), a key intervention recommended by the WHO to achieve their End TB Strategy targets [1]. The efficacy of current WHO-recommended TB preventive treatment for drug-sensitive TB ranges from 60% to 90% [2]. Studies have demonstrated the effectiveness of TPT among high-risk groups, including people living with HIV (PLHIV), those experiencing homelessness, close contacts of pulmonary TB cases (children and adults), and immunocompromised people [3]. Significant progress has been made in improving TPT for those exposed to drug-sensitive TB. Current WHO recommendations include a shorter isoniazid-rifapentine based regimen (ranging from one to three months) with higher treatment completion rates and significantly less adverse drug reactions (ADR) [3,4].

While the expansion of newer TPT regimens for exposure to drug-sensitive strains of TB has helped scale up the search-treat-prevent strategy to eliminate TB [5], it does not address how to treat contacts exposed to drug-resistant strains of *M. tuberculosis* (DR-TB). To date, DR-TB remains a significant contributor to global antimicrobial resistance, and untreated exposure to drug-resistant strains continues to cause disease, ranging from disease caused by isoniazid and rifampin mono-resistant (RR-TB) to multi- and extensively drug-resistant strains (MDR-TB and XDR-TB). A meta-analysis of 25 studies found that 47.2% of household contacts of MDR-TB patients were diagnosed with TB infection, and 7.8% of household contacts developed TB, most within one year of the source case’s diagnosis [6]. Although the WHO recommended TPT for high-risk household contacts of patients with multidrug-resistant tuberculosis (based on individualized risk assessments) in 2018, and strengthened this recommendation in 2024 to include 6 months of daily levofloxacin as TPT, such care remains neither widespread nor mainstream [3,7]. This is particularly important in the 30 high-MDR/RR-TB-prevalence countries, including China, India, Indonesia, Pakistan, Philippines, Russia, South Africa, and Viet Nam [8].

Significant gaps remain in determining optimal TPT regimens for those exposed to DR-TB strains. A systematic review of TPT for TB infection caused by DR-TB showed the effectiveness of fluoroquinolone-containing regimens [9]. In an analysis of 12 studies, the average proportion of individuals experiencing ADRs resulting in treatment discontinuation was 19% (106/558; 95% CI, 16%–22%). Fluoroquinolone (FQ)–containing regimens (without pyrazinamide) had a high percentage (33%) of adverse events but rarely led to discontinuation (only 2% of the time). A cohort studying Arkhangelsk, Russian Federation, shows that MDR-TB TPT with FQ is safe and well-tolerated by children [10].

However, a significant proportion of MDR-TB globally is fluoroquinolone resistant (19% in 2023) [11], and no recommendations exist for TPT for close contacts of people infected with these strains. With this in mind, Vladimir province in the Russian Federation turned to bedaquiline, a newer TB drug, as TPT for such individuals. Bedaquiline is highly bactericidal and effective against both drug-sensitive and drug-resistant strains, including fluoroquinolone-resistant strains [12]. In a murine model of tuberculosis infection, bedaquiline effectively treated drug-resistant TB with sterilizing activity of not less than rifapentine [13]. In clinical trials, most adverse events were mild to moderate [12].

In this study, we aimed to assess the safety, efficacy and feasibility of a comprehensive TPT program among several risk groups in Vladimir City, including contacts of individuals with drug-sensitive and drug-resistant TB, including fluoroquinolone-resistant forms. Our specific objectives were to determine: 1) The proportion of ADR type and severity (grade) by TPT regimen; 2) The rate of TPT completion by regimen, ADRs and DOT type; and 3) TB incidence among enrolled patients in the 12-month follow-up.

## METHODS

### Setting and Study Design

We conducted a retrospective cohort study as part of the Zero TB Initiative in Vladimir City, Russia. Age and gender distribution of the study population align with national averages (adults 16-54 comprise 62% of the population with a slight predominance of women, a common feature in Russia) [14]. About 11.8% of residents live below the poverty line in 2021 [15]. TB and DR-TB are managed by the Vladimir Regional TB Control Center through inpatient and outpatient facilities in regional centers, with no private sector involvement. TB/HIV cases are jointly managed with the Regional Center for AIDS Prevention.

According to the Federal Center for Monitoring Tuberculosis Control in Russia (Federal Statistical Monitoring Form No. 8), there has been a steady, systematic decline in TB incidence in Vladimir Oblast, from 50.6 per 100,000 people in 2014 to 36.0 per 100,000 people in 2018 [16]. Meanwhile, the proportion of primary MDR-TB remained stable: 22.1% in 2014 to 22.6% in 2018 (Form No. 7-TB) [17]. Additionally, Federal Statistical Monitoring Form No. 33 reported that the proportion of HIV-positive individuals among newly identified TB cases rose by 2.3% since 2014, reaching 11.0% in 2018.

At the end of 2018, the Zero TB Initiative was launched in Vladimir City to reduce tuberculosis rates in the region [18]. As part of this initiative [5], a comprehensive search-treat-prevent program for TB elimination was initiated, including TPT delivery to high-risk groups. We analyzed data collected from this initiative. The study protocol was adapted from a freely available handbook [19].

### Study Population and Enrollment

We retrospectively analyzed data from individuals over 18 who belonged to risk groups (close contacts of individuals with drug-sensitive and DR-TB, people experiencing homelessness, and people living with HIV (PLHIV)) and received TPT between January 1, 2019, and December 31, 2020 in the civilian sector of Vladimir city.

Persons experiencing homelessness and PLHIV were eligible for TPT regardless of known TB contacts. Children under 18 were excluded as were people denied of liberty. All individuals were screened as part of routine program implementation using a predetermined screening algorithm for TB, which included symptom evaluation, chest X-ray, and Diaskintest [20, 21]. HIV testing was recorded for those with unknown status, and Xpert MTB/RIF assay was performed if sputum was produced.

The program prioritized care for household contacts of TB patients and close non-household contacts with at least 8 h/day or 12 h/week of exposure; all others were considered lower priority. Contact tracing varied by index case characteristics: for sputum smear-positive cavitary/laryngeal TB, both high- and low-priority contacts were monitored; for sputum smear-negative, non-cavitary pulmonary TB, only high-priority contacts were followed.

### Study Intervention

Individuals diagnosed with active TB were recorded as enrolled in the TB treatment program. Those without active TB but with a positive Diaskintest (indicated by erythema or induration/papule of any size) were eligible for TPT based on programmatic criteria. TPT was also provided to PLHIV and high-priority contacts, regardless of their Diaskintest result. Individuals not meeting these criteria were monitored for potential development of active TB, but did not receive TPT. People exposed to drug-sensitive TB received a rifapentine/isoniazid regimen (3HP). In cases of contraindications or drug interactions, alternative regimens—rifabutin/isoniazid (3HRb) or isoniazid alone (6H)—were used. Those exposed to isoniazid-resistant TB received a rifampicin regimen (4R), and those exposed to rifampicin-resistant TB were given moxifloxacin (4Mfx). For individuals exposed to strains resistant to both rifampicin and fluoroquinolones, bedaquiline (3Bdq) was prescribed (Table 1).

**Table 1.**
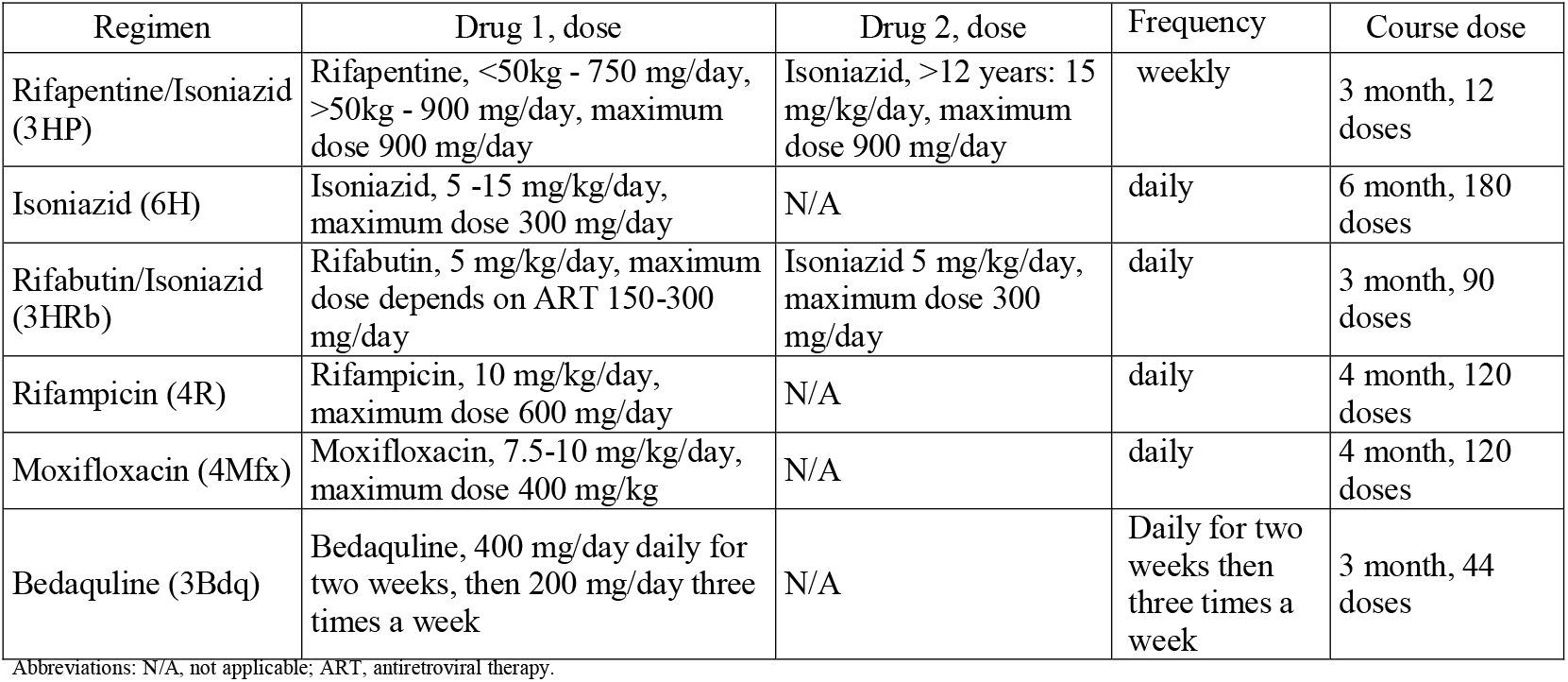
TPT regimens used in the study among PLHIV, homeless people and contact persons, Vladimir, Russia, January 2019 – December 2020.

As part of the Zero TB Initiative, medical staff had been trained on TB screening and prevention, and educational leaflets were developed for TB patients and their contacts. To reduce barriers to care, household screening was conducted, and all TB screening procedures were completed in a single clinic visit with flexible scheduling. Occupational contacts were screened at their workplaces. Psychological counseling was made available to patients and their relatives. Directly observed therapy (DOT) was tailored to patient preferences, and all services, including screening, diagnosis, TB treatment, TPT, and access to side-effect medications, were free. Patients and medical staff received financial incentives (approximately $5 USD) for completing each step of the care cascade.

TPT was primarily supervised by medical personnel using video-supported treatment (VST), a mobile team, or medication administration in the outpatient treatment room at either the TB or HIV care facilities. Patients using VST or their caregivers had mobile devices to connect with medical staff or send video proof of medicine intake. Most 3HP patients received treatment at outpatient facilities, while mobile teams served those unable to use VST or visit clinics, including homeless individuals. Self-administered treatment (SAT) was permitted for patients working in remote areas without access to mobile service and/or internet, with adherence monitored through monthly clinic visits for prescriptions and follow-up exams.

During monthly follow-up visits, patients underwent TB symptom screening, physical examination, ECG, liver function tests, blood counts, and metabolic panels as part of routine program monitoring. Patients with symptoms were examined weekly by a physician. Adverse drug reactions (ADRs) were graded according to the Common Terminology Criteria for Adverse Events (CTCAE), version 5.0 [22].

A committee of TB physicians determined appropriate interventions for patients with ADRs. The most common ADRs were flu-like syndrome, hepatic dysfunction (any increase in liver enzymes such as ALT (alanine aminotransferase) or AST (aspartate aminotransferase) or bilirubin above the upper limit of normal), hepatotoxicity (a threefold elevation in AST or ALT above the upper limit of normal (ULN)) and vomiting. Post-ADR outcomes included treatment continuation, switching regimens, or patient/doctor-initiated treatment termination.

If a patient was febrile (flu-like syndrome), the regimen was discontinued or switched, regardless of adverse reaction grade, and the patient was closely followed. In the case of hepatic dysfunction, TPT was temporarily stopped for transaminase levels two times the upper normal limit, and patients were followed for 4-6 weeks. Usually a course of hepatoprotective agents, mostly ademetionine, was prescribed. If transaminases dropped to normal levels, TPT was continued; if not, TPT was terminated.

Treatment was classified as ‘completed’ if at least 85% of the prescribed doses were taken and the TPT interruption period was less than 2 weeks; anything below this threshold was considered ‘non-completion”. Non-completion was further categorized as due to adverse drug reactions and loss to follow-up. All individuals were observed for at least 12 months, with follow-up chest x-rays to assess for TB. However, not all individuals attended the follow-up chest x-ray on time, so the follow-up radiological examination was accepted between 12 and 24 months.

### Analysis

Data used for this analysis was extracted from patient registration forms and double-entered into an Access Data Base at the Vladimir Zero TB project. Descriptive statistics summarized patient characteristics, completion rates of each TB infection cascade step, and reasons for discontinuation. Outcome measures included adverse events, TPT completion, and TB disease incidence during the 12-month follow-up period. We accounted for differing times of the final one-year follow-up x-ray with an analysis of person years of observation.

We assessed safety by calculating the proportion of persons with ADRs based on TPT regimen and sex; analyzed ADR types and severity; treatment completion rates. Feasibility of TPT delivery was assessed by tracing participants through the steps of the care cascade (evaluation for TB, offer TPT, initiation, completion, and 12-month follow-up; see Figure 1) and calculating completion rates for each step, cross-tabulated by regimen and DOT type.

**Figure 1.**
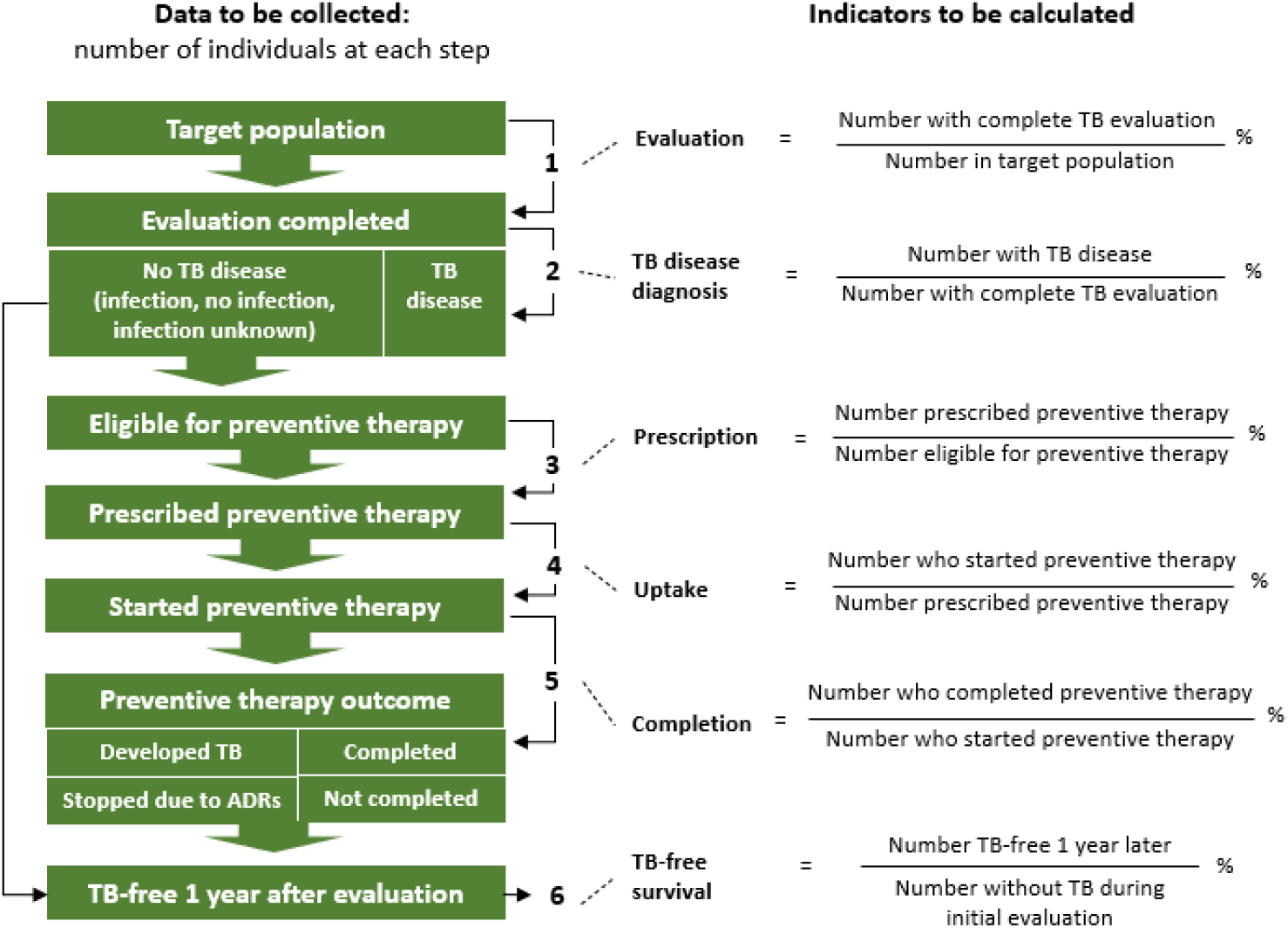
TB prevention cascade. ^i^ The Zero TB project in the city of Vladimir, under which operational research was conducted, was funded through a grant from the Eli Lilly and Company Foundation to Brigham and Women’s Hospital (USA) (S.K. was the Boston-based principal investigator). Data collection according to standards for the study and data analysis was supported by Partners in Health, Boston, USA. The preparation of the first draft of the manuscript was initiated during the SORT IT course funded by “LHL international”, Norway (A.V.S. attended this course in 2021). ^ii^ The authors have declared that no competing interests exist.

Statistical analysis was performed using OpenEpi (Version 3) and Microsoft Excel. χ^2^ and Mid-P exact tests were used to assess differences between groups, with significance set at p < 0.05. Odds ratios (ORs) and confidence intervals (CIs) were calculated for variables with significant differences.

### Ethical Approval

Institutional Review Board approval was obtained in Russia and the United States. All participants signed an informed consent at the stage of treatment enrollment with the Vladimir TB services.

#### RESULTS

Between January 2019 and December 2020, 3,822 representatives of target groups were screened for TB using the project algorithm. At baseline evaluation, active TB disease was diagnosed in 24 individuals (0.6%). Of the remaining 3,798 individuals, 606 (16.0%) were eligible for TPT, and it was offered to 77.1% of them (467 out of 606). Known reasons for not offering TPT included change of residence, medical contraindications, and COVID lockdown. Of these, 50 individuals (10.7%) declined the offer, and 14 (3.0%) accepted but did not start TPT.

Table 2 describes demographics and clinical characteristics of the study population at enrollment.

**Table 2.** Demographics and clinical characteristics of study population free of TB disease at baseline in study of preventive therapy in Vladimir, Russia, January 2019 – December 2020.

| Characteristics | Total, n (%) or median [IQR] | Eligible and initiated TPT, n (%) or median [IQR] | Eligible but did not initiate TPT, n (%) or median [IQR] | Not eligible for TPT, n (%) or median [IQR] |
| --- | --- | --- | --- | --- |
|  | (N = 3798) | (n = 403) | (n = 203) | (n = 3192) |
| Age, y | 40.7 [31.6-51.7] | 41.7 [34.7-51.3] | 42.5 [35.3-51.0] | 40.4 [30.8-51.8] |
| Sex (male) | 1949 (51.32) | 234 (58.1) | 122 (60.1) | 1593 (49.9) |
| Presence of TB symptoms or pathological chest X-ray (CXR) | 190 (5.0) | 61 (15.1) | 32 (15.8) | 97 (3.0) |
| Known TB contact | 2969 (78.2) | 149 (37.0) | 59 (29.1) | 2761 (86.5) |
| HIV-positive status | 308 (8.1) | 195 (48.4) | 112 (55.2) | 1 (0.03) |
| Homelessness | 607 (16.0) | 78 (19.4) | 67 (33.0) | 462 (14.5) |
| Positive Diaskintest result | 285 (7.5) | 201 (49.9) | 84 (41.4) | 0 (0) |
| Drug use | 62 (1.6) | 25 (6.2) | 16 (7.9) | 21 (0.7) |
| Harmful alcohol consumption | 437 (11.5) | 66 (16.4) | 46 (22.7) | 325 (10.2) |
| Other known comorbidities | 142 (3.7) | 70 (17.4) | 34 (16.7) | 38 (1.2) |
| Regimen given: |  |  |  |  |
| 3HP | N/A | 253 (62.8) | N/A | N/A |
| 3HRb | N/A | 23 (5.7) | N/A | N/A |
| 6H | N/A | 55 (13.6) | N/A | N/A |
| 4R | N/A | 22 (5.5) | N/A | N/A |
| 4Mfx | N/A | 29 (7.2) | N/A | N/A |
| 3Bdq | N/A | 21 (5.2) | N/A | N/A |
| Follow-up CXR was done | 1903 (50.1) | 294 (73.0) | 129 (63.5) | 1480 (46.3) |
| TB disease occurred during follow-up period in individuals who initiated treatment and completed TPT | N/A | 0 (0) | N/A | N/A |
| TB disease occurred during follow-up period in individuals who initiated treatment but did not complete TPT | 5 (0.1) | 1 (0.25) | 2 (1.0) | 2 (0.06) |
| Rates of TB disease per 1000-person years of observation<sup>1</sup>, n [95% CI] | N/A | 2.9 [2.7, 3.08] | 11.6 [11.1, 12.1] | 1.05 [1.048, 1.051] |

Abbreviations: IQR, interquartile range; N/A, not applicable; Observation period is defined as the period from the first chest x-ray (CXR) to the final x-ray at the end of the 12-month follow-up period. Since some patients could not come for their CXR at exactly the 12-month mark, CXR results up to 24 months were accepted as an indication of the 12-month follow-up period. Individuals without a follow-up CXR (no CXR before month 24) were not included in the person-years of observation; 95% CI, confidence interval.

Adverse drug reactions (ADRs) were recorded in 67 out of the 403 (16.6%) individuals who started TPT (Table 3). Forty-six (11.4%) individuals reported at least one ADR during treatment and 21 (5.2%) reported more than one ADR. Forty-five (67.2%) of the 67 participants who experienced ADRs discontinued treatment.

**Table 3.** Number of persons reported any adverse drug reaction (ADR) among those who initiated tuberculosis preventive treatment (TPT) by sex and regimen in Vladimir, Russia, January 2019 – December 2020.

| Variables | Started TPT,<br>n | Reported ADR,<br>n (%) | P-value | OR | 95% CI |
| --- | --- | --- | --- | --- | --- |
| Total | 403 | 67 (16.6) |  |  |  |
| Sex: |  |  |  |  |  |
| Male | 234 | 27 (11.6) | 0.002<sup>1</sup> | 0.42 | 0.25, 0.72 |
| Female | 169 | 40 (23.7) |  |  |  |
| TPT regimens: |  |  |  |  |  |
| 3HP | 253 | 37 (14.6) | Ref. |  |  |
| 3HRb | 23 | 10 (43.5) | 0.002<sup>2</sup> | 4.5 | 1.77, 11.02 |
| 4R | 22 | 2 (9.1) | 0.5<sup>2</sup> |  |  |
| 6H | 55 | 8 (14.5) | 0.8<sup>1</sup> |  |  |
| 4Mfx | 29 | 9 (31.0) | 0.04<sup>2</sup> | 2.6 | 1.06, 6.14 |
| 3Bdq | 21 | 1 (4.8) | 0.2<sup>2</sup> |  |  |

<sup>1</sup>, Yates corrected chi square; Ref., reference data; <sup>2</sup>, Mid-P exact.

Table 4 shows 94 ADRs reported from 67 individuals undergoing preventive treatment. No life-threatening ADRs or deaths were reported. Hepatic dysfunction was the most common for the 4Mfx group, but all 5 cases were Grade 1 severity and did not lead to treatment discontinuation. The 3HP-associated flu-like syndrome typically occurred after a median of three doses (3 weeks) [21—23], while in the 3HRb group, it appeared after a median of seven doses (7 days). The lowest rate of ADRs was observed in 3Bdq group, reported in one out of 21 patients on TPT. This patient, who had taken 75% of the doses, also experienced diabetes decompensation. No QT prolongation was observed.

**Table 4.** Number, type and severity of adverse drug reactions experienced during TB preventive treatment, by regimen in Vladimir, Russia, January 2019 – December 2020.

| Averse drug reactions | Total number of ADRs, n (%) | 3HP, n (%) | 3HRb, n (%) | 4R, n (%) | 6H, n (%) | 4Mfx, n (%) | 3Bdq, n (%) |
| --- | --- | --- | --- | --- | --- | --- | --- |
|  | (N = 94) | (n = 58) | (n = 11) | (n = 3) | (n = 11) | (n = 9) | (n = 2) |
| Severity: |  |  |  |  |  |  |  |
| Grade I | 45 (47.9) | 26 (44.8) | 8 (72.7) | 1 (33.3) | 4 (36.4) | 6 (66.7) | 0 (0.0) |
| Grade II | 32 (34.0) | 22 (37.9) | 1 (9.1) | 2 (66.7) | 5 (45.5) | 1 (11.1) | 1 (50.0) |
| Grade III | 17 (18.1) | 10 (17.2) | 2 (18.2) | 0 (0) | 2 (18.2) | 2 (22.2) | 1 (50.0) |
| Type: |  |  |  |  |  |  |  |
| Flu-like syndrome | 22 (23.4) | 15 (25.9) | 7 (63.6) | 0 (0) | 0 (0) | 0 (0) | 0 (0) |
| Hepatic dysfunction | 13 (7.2) | 7 (12.1) | 0 (0) | 0 (0) | 1 (9.0) | 5 (55.6) | 0 (0) |
| Hepatotoxicity | 19 (20.2) | 11 (19.0) | 3 (27.3) | 1 (33.3) | 2 (18.2) | 1 (11.1) | 1 (50.0) |
| Vomit/appetite | 13 (13.8) | 7 (12.1) | 1 (9.1) | 1 (33.3) | 3 (27.3) | 0 (0.0) | 1 (50.0) |
| Others ADRs | 27 (28.7) | 18 (31.0) | 0 (0) | 1 (33.3) | 5 (45.5) | 3 (33.3) | 0 (0) |

ADRs, adverse drug reactions; Hepatic dysfunction, any increase in ALT, AST, or bilirubin above the upper limits of normal; Hepatotoxicity, a 3 times elevation in ALT or AST above the upper limit of normal (ULN); Others ADRs: allergy, diarrhea, headache, high blood pressure, pancreatitis, psychosis, rash, syncope, weakness/irritability.

Treatment completion rates ranged from 52.2% to 95.2%. The probability of completing treatment if TPT was offered was 64.5% (301 of 467). The completion rate varied by TPT regimen and was significantly higher for 3Bdq (n=20, 95.2%) compared to 3HP (n=192, 75.9%, *Mid-P exact* = .03). The non-completion rate was significantly higher for 3HRb (n=11, 47.8%) compared to 3HP (n=61, 24.1%, χ ^2^ *P* value = .01). Among DOT types, outpatient treatment clinics (50%) and video-supported treatment (31%) were more common. The completion rate was lower among individuals on self-administered treatment (n=19, 59.4%, χ^2^ P-value = .03) and those receiving TPT administration via mobile teams (n=26, 59.1%, χ^2^ P-value = .01) compared to those receiving TPT in outpatient clinics (n=159, 79.1%).

Over the 12-month follow-up, two of the 403 individuals who began TPT died from causes not related to TB. Chest X-ray coverage was 79.4% among those who completed the full TPT course, compared to 61.3% in individuals who were eligible but either did not start or complete treatment. No active TB disease occurred in those who completed the full course of TPT. One case occurred in an individual who started TPT but did not complete treatment (he had only taken 2 doses of 3HP, experienced no ADRs, and developed drug-sensitive TB at the 9-month mark). One case occurred in an individual who was eligible but did not start TPT. The rate of TB disease per 1,000 person-years of observation was four times higher in individuals eligible for TPT who did not start it compared to those who initiated TPT (Table 2).

## DISSCUSSION

This analysis demonstrates that providing TPT for individuals exposed to drug-sensitive and DR-TB strains is feasible and safe, including those resistant to rifampicin and fluoroquinolones. All six regimens were well tolerated, with the highest completion rate and lowest adverse events observed in the 3Bdq group. No TB cases were reported during the 12-month follow-up among those who completed TPT. Moreover, the protective effect of TPT was 75%, consistent with findings from two studies on Isoniazid for TPT [24, 25]. To our knowledge, this is the first study to offer a bedaquiline-based preventive therapy to contacts of fluoroquinolone-resistant TB (pre-XDR TB) in a high MDR-TB burden setting, and the first report of ADRs for TPT using bedaquiline.

Treatment termination due to ADRs occurred in 11%; other studies range from 1.4% to 17% [26— 28]. The proportion of individuals experiencing ADRs on the 4Mfx regimen that led to treatment discontinuation was 6.9%, which is lower than reported in a systematic review where 43 reported ADRs led to 4 (9.3%) discontinuations on fluoroquinolone regimen [9]. The completion rate for 4Mfx in our study was 87%, matching the 89% reported in a study from the Federated States of Micronesia [29], but higher than the completion rate of 68% reported in Karachi [30].

Other studies found no evidence that DOT had better cure or completion rates than SAT, suggesting treatment duration impacts completion rates more than monitoring mode [31—Click or tap here to enter text.33]. This aligns with our study, which showed the highest non-completion rates for TPT organized via mobile teams and SAT, primarily due to loss to follow-up. Of those who discontinued, 89% were persons experiencing homelessness, and 61% had alcohol use disorder. The low completion rate of 64% among persons experienced homelessness highlights their vulnerability to not completing long TPT courses. A shorter regimen, as recommended by WHO [34], may improve outcomes.

Key strengths of this analysis included its rigorous programmatic design and evaluation, the use of directly observed TPT in most cases, a strong system for detecting and managing adverse drug reactions, and the integration of financial incentives to help mitigate poverty-related barriers to treatment adherence. Our study had several limitations. The inclusion of a broad range of occupational contacts, including low-priority groups, may have contributed to the low baseline TB prevalence among contacts. Of the 3,822 participants, 78% were contacts, and 92% (2,732/2,969) of those screened were occupational contacts. Among occupational contacts, 94% (2,564/2,732) were ineligible for preventive therapy. No children were included, and some groups were small due to regimen assignment based on TB strain exposure. Additionally, some patients were lost to follow-up at the 12-month mark. However, TB treatment was only available through the governmental system, facilitating tracking of new TB cases within the cohort. All TB cases in Russia are registered in a federal TB registry, enabling follow-up even if a patient relocated.

## Conclusion

This study is the first to assess the safety, efficacy and feasibility of bedaquiline for tuberculosis preventive treatment. It introduces a novel paradigm for TB prevention in high-burden pre-XDR-TB settings, offering a promising strategy to protect close contacts and reduce TB transmission. We found that providing TB preventive treatment for people infected with drug sensitive and drug-resistant strains of TB is both feasible and safe, even for those exposed to strains resistant to rifampicin and fluoroquinolones. No TB disease was observed during the 12-month follow-up period among individuals who completed the full course of TPT. The highest TPT completion rates and lowest adverse events rates were observed in the 3Bdq group, suggesting that a larger study on the use of Bdq-containing regimens for treatment of tuberculosis infection is warranted.

## Supporting information

Figure 1. TB prevention cascade.

## Data Availability

All data produced in the present work are contained in the manuscript

## Notes

### Author contributions

A. V. S., G. V. V, S. K., S. G. H. conceptualized the study and wrote the protocol. T. A. K., T. R. S,, E. V. B., A. V. S. implemented the study and collected data under supervision from G. V. V, O. I. P. and S. K.. A. V. S., and S. G. H., S. K., E.H. performed and reviewed the analysis. A. V. S., S. K., wrote the initial draft of the manuscript. All authors helped interpret the findings and read and approved of the final version of the manuscript.

## Acknowledgments

Special thanks to: Elena S. Dyuzhik, Ekaterina A. Volkova, and all the staff of the Vladimir Regional TB Control Center; Svetlana N. Makarova, and all the staff of the Regional Center for AIDS Prevention who worked to implement the comprehensive search-treat-prevent program for TB elimination; Nina V. Kuteneva and all the staff of the Russian Independent Nonprofit Organization Center of Partnership Assistance in Healthcare “Zdorovye.ru” who provided technical assistance for the program’s implementation; Drs. Andrei Mariandyshev, as a co-organizer and tutor of the SORT IT (Structured Operational Research and Training Initiative) course in Arkhangelsk city in 2021–2022, who provided feedback on the protocol draft and analysis; and for comments on the manuscript. Data not publicly available.

## Disclaimer

The funder of this study had no role in study design, data collection, data analysis, data interpretation, writing of the report, or in the decision to submit for publication. The authors had full access to the data and made the decision to publish the manuscript.

