## Supplementary material for "Safety, efficacy and feasibility of preventive treatment for drug-resistant tuberculosis with moxifloxacin or bedaquiline": Figure 1. TB prevention cascade.: Figure_1.pdf

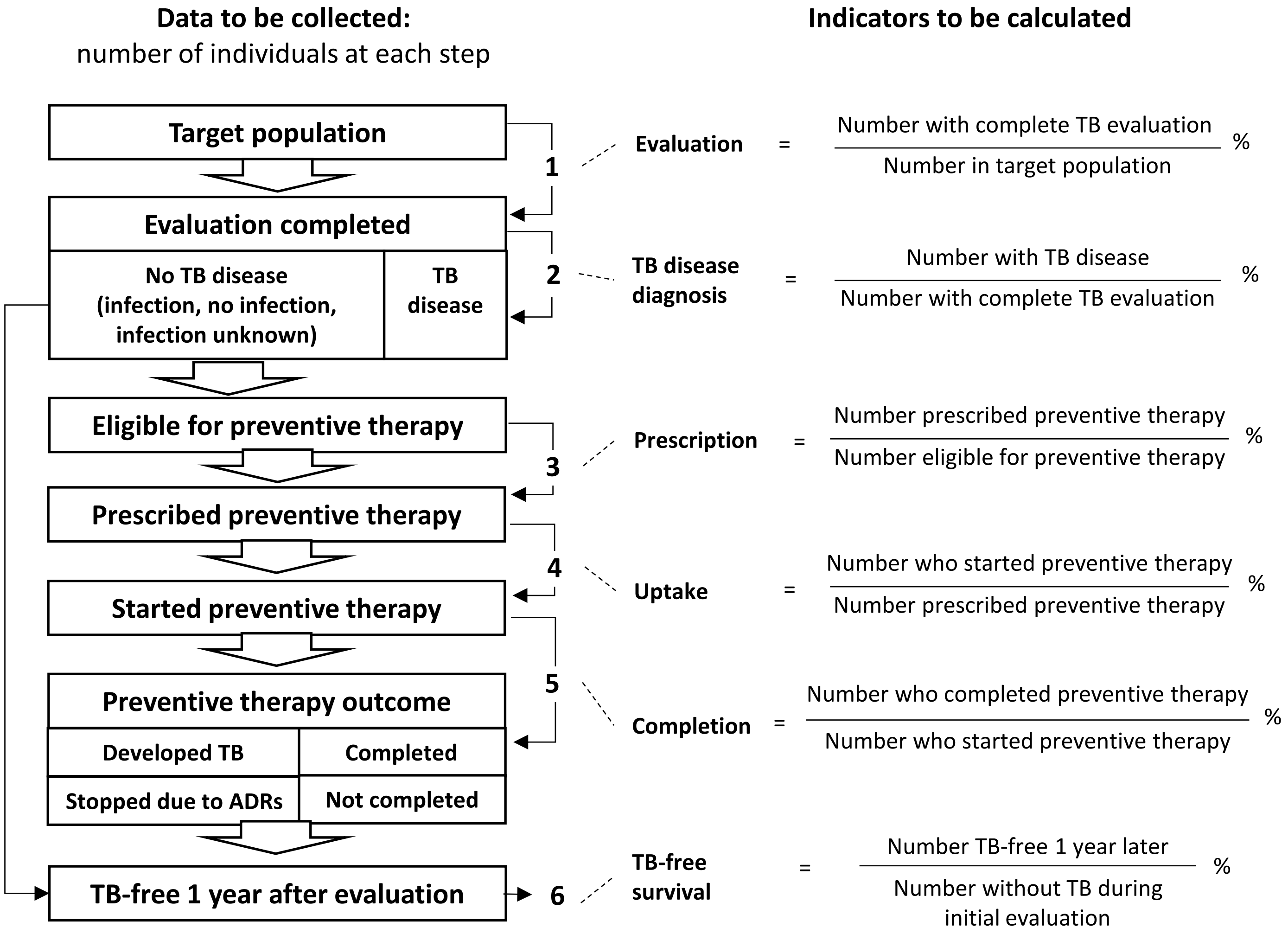
Data to be collected:
number of individuals at each step
Indicators to be calculated
Target population
Evaluation completed
No TB disease (infection, no infection, infection unknown)
TB disease
Eligible for preventive therapy
Prescribed preventive therapy
Started preventive therapy
Preventive therapy outcome
Developed TB
Completed
Stopped due to ADRs
Not completed
TB-free 1 year after evaluation
1
2
3
4
5
6
Evaluation = Number with complete TB evaluation / Number in target population %
TB disease diagnosis = Number with TB disease / Number with complete TB evaluation %
Prescription = Number prescribed preventive therapy / Number eligible for preventive therapy %
Uptake = Number who started preventive therapy / Number prescribed preventive therapy %
Completion = Number who completed preventive therapy / Number who started preventive therapy %
TB-free survival = Number TB-free 1 year later / Number without TB during initial evaluation %

**Data to be collected:**
number of individuals at each step

**Indicators to be calculated**

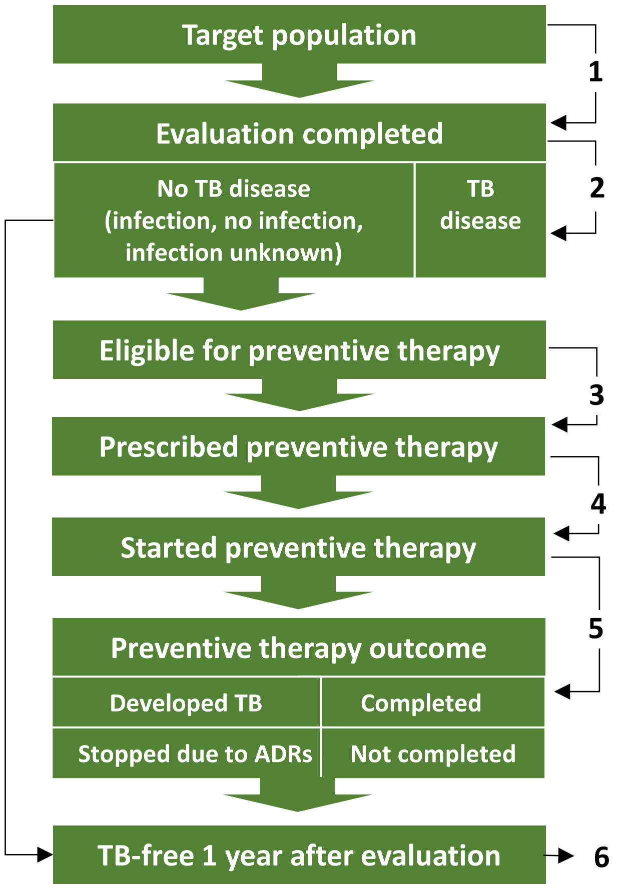


1. $$\textbf{Evaluation} = \frac{\text{Number with complete TB evaluation}}{\text{Number in target population}}\ \%$$

2. $$\textbf{TB disease diagnosis} = \frac{\text{Number with TB disease}}{\text{Number with complete TB evaluation}}\ \%$$

3. $$\textbf{Prescription} = \frac{\text{Number prescribed preventive therapy}}{\text{Number eligible for preventive therapy}}\ \%$$

4. $$\textbf{Uptake} = \frac{\text{Number who started preventive therapy}}{\text{Number prescribed preventive therapy}}\ \%$$

5. $$\textbf{Completion} = \frac{\text{Number who completed preventive therapy}}{\text{Number who started preventive therapy}}\ \%$$

6. $$\textbf{TB-free survival} = \frac{\text{Number TB-free 1 year later}}{\text{Number without TB during initial evaluation}}\ \%$$
